# Which innovations can improve timeliness of investigations and address the backlog in endoscopy for patients with potential symptoms of upper and lower Gastrointestinal (GI) cancers?

**DOI:** 10.1101/2022.05.04.22274653

**Authors:** Annie Hendry, Llinos Haf Spencer, Ned Hartfiel, Bethany Anthony, Jessica Roberts, Joanna M Charles, Nathan Bray, Clare Wilkinson, Rhiannon Tudor Edwards

## Abstract

**What is a Rapid Review?:** Our rapid reviews use a variation of the systematic review approach, abbreviating or omitting some components to generate the evidence to inform stakeholders promptly whilst maintaining attention to bias. They follow the methodological recommendations and minimum standards for conducting and reporting rapid reviews, including a structured protocol, systematic search, screening, data extraction, critical appraisal, and evidence synthesis to answer a specific question and identify key research gaps. They take 1-2 months, depending on the breadth and complexity of the research topic/ question(s), extent of the evidence base, and type of analysis required for synthesis.

**Background / Aim of Rapid Review:** Many patients were not able to access routine diagnostic care through 2020/21 because of extraordinary pressures on the NHS due to COVID-19 and the UK national lockdowns. For some patients this can have serious short and long-term consequences to their health and life expectancy. The NHS has limited resources and is looking for new ways to meet many demands and patient needs.

This Rapid Review Report aims to answer the question “Which innovations can be used to accelerate the patients’ journey through the endoscopic cancer diagnosis pathway?” The report highlights evidence of innovations and new ways to improve the timeliness of access to endoscopy and to address the backlog of unmet need for patients who have waited a long time for such tests and investigations by selecting those at highest for prioritisation. It does not evaluate in terms of effectiveness on clinical outcomes.

**Key Findings:** *Extent of the evidence base:*

▪ Nine papers were included in the rapid review in total.
▪ Two reviews were identified. One review examined the novel **colon capsule endoscopy (CCE)** procedure and the second review summarised the effects of COVID-19 on colorectal cancer (CRC) screening, the potential long-term? outcomes, and ways to adapt CRC screening during the COVID-19 pandemic.
▪ Seven primary studies assessed innovations for the diagnosis of Gastrointestinal (GI) cancers. Five of these studies examined **faecal immunochemical testing (FIT)** for prioritising patients for further testing.
▪ Two studies reported **pathways/innovations** to triage patients e.g. from primary care. These methods of triage used interventions such as Cytosponge for oesophageal symptoms.

*Recency of the evidence base:*

▪ Of the primary studies, one was published in 2020 and six were published in 2021. Of the reviews, one was published in 2020 and one in 2021.

*Evidence of effectiveness:*

▪ The five studies investigating FIT found that it could help prioritise patients for further testing and improve targeting of high-risk patients.
▪ One review proposed CCE may offer a useful solution for investigating colorectal patients to reduce the need for some endoscopies following the pandemic.
▪ One review found a shift from current CRC screening and surveillance practices towards an individualized approach based on risk factors, could result in the allocation of resources to people with higher risks and prevent inappropriate use of healthcare resources for those with lower risks.

*Best quality evidence:*

▪ All studies were quality appraised using the relevant JBI checklist. Five studies were of low to moderate quality.

**Policy Implications:**

▪ Increased use of faecal immunochemical testing (FIT) could reduce the endoscopy backlog and save NHS resources if those with low FIT scores can be excluded from further testing.
▪ Policy in Wales supports prioritisation of potential gastrointestinal cancer patients for endoscopy using FIT test scores (NHS Wales 2021) although local implementation currently varies, so it is not yet fully utilised. The FIT test gives results which could be utilised by healthcare professionals to prioritise those who are most in need of urgent diagnosis. The viability of this method to prioritise those in greatest need of being referred for diagnosis through endoscopy is proven (though safety-netting is still required), and the FIT test is part of the diagnostic pathway already in Wales. It will be important to ensure all areas of Wales have equal access to the use of FIT testing for this purpose, and that clinical guidelines are harmonised and adhered to throughout Wales.
▪ Innovations to reduce backlog and speed up time to diagnosis should be explored including:
  ○ Triage in primary care settings such as GP surgeries using innovations such as the cytosponge for oesophageal symptoms (e.g. reflux).
  ○ Direct referral from primary care settings to specialist investigation, without the need for prior additional referrals in secondary care.

**Strength of Evidence:**

▪ The evidence presented in this review is recent, however with small samples (di Pietro et al., 2020), short-term follow up periods (Sagar et al., 2020) and assumptions required for modelling studies (Loveday et al., 2021). This reduces the generalisability and confidence of conclusions. The confidence in the strength of evidence about FIT testing is rated as ‘low-moderate confidence’. Cytosponge evidence is rated ‘low confidence’.

**Review team and stakeholder involvement:** This Rapid Review is being conducted as part of the Wales COVID-19 Evidence Centre Work Programme. The above question was developed in consultation with Cancer Research UK’s identified research gaps and with Professor Tom Crosby OBE. Professor Crosby is a Consultant Oncologist, National Cancer Clinical Director for Wales and Clinical Lead for Transforming Cancer Services and acted as the expert stakeholder for this review.

The search questions were identified as a priority during the Cancer/COVID-19 Research Summit hosted by Cancer Research UK (CRUK), Public Health England (PHE) and the National Cancer Research Institute (NCRI). The stakeholder group supporting the review work here is Cancer Research Wales.

## 1. BACKGROUND

Many patients were not able to access routine diagnostic care through 2020/21 due to extraordinary pressures on the NHS due to the COVID-19 pandemic restrictions and the UK national lockdowns. For some patients this had serious short and long-term consequences to their health and life expectancy. The NHS has limited resources and is looking for new ways to meet many demands and patient needs.

There are approximately 2,200 cases of colon cancer diagnosed in Wales each year (Bowel Cancer UK, 2018). In 2018, there were 1,610 patients waiting over the 8-week diagnostic target for endoscopy services in Wales (National Assembly for Wales, 2019). In the interest of clarity, the term ‘endoscopy’ is used in this review as an overarching term for procedures where a small camera inserted into the body to examine internal organs, including colonoscopy (looking at the colon and rectum) and gastroscopy (looking at the oesophagus and stomach).

Nodora et al. (2020) noted that the COVID-19 pandemic may further exacerbate existing colorectal cancer (CRC) screening disparities in underserved individuals which will likely lead to delayed diagnosis, a shift to later stage disease, and potentially also increased CRC deaths (Forbes et al, 2021). To prevent this from happening, Nodora et al. (2020) call for timely action and a commitment to address the current extraordinary CRC screening challenges for vulnerable populations.

Das (2020) conducted a Discrete Event Simulation (DES)–based model study in the setting of a small to medium community-based single-specialty ambulatory endoscopy centre (AEC). This study quantified the impact of COVID-19 related workflow changes on performance indicators and cost per case compared with the pre–COVID-19 baseline. DES-based modelling is a novel method for formal quantitative assessment of resource use, throughput, and capacity constraints of complex systems by simulating dynamic interactions between individuals, populations, and their environments using a sequence of well-defined events and focusing on individual entities (e.g., patients) moving through the system with changes in their health states at discrete time points. Pretesting and screening for COVID-19 as recommended by current guidelines significantly influenced the productivity and revenue stream of ambulatory endoscopy centres. Das, (2020) noted that urgent measures by payers are needed to adjust the facility reimbursement of endoscopy centres to ensure successful reopening and ramping up outpatient endoscopy services in these facilities already hit hard by the pandemic.

Interventions to reduce endoscopy or prioritise referrals include cytosponge testing and faecal immunochemical testing (FIT). The Cytosponge is a non-endoscopic diagnostic tool consisting of a tethered capsule, which is swallowed and collects immunohistochemical biomarkers of intestinal metaplasia and dysplasia. Cytosponge can be administered in primary (reflux symptoms) or secondary care (e.g. as part of Barrett’s surveillance) settings. FIT is a test designed to identify blood in a person’s stool, which may be a sign of colorectal cancer (CRC).

FIT tests were adopted in the UK in 2019 and can be used for both screening and for prioritising of symptomatic patients. Data from this study was used to create a prioritisation model for endoscopy and 2WW referrals during the COVID-19 pandemic.

The initial searches were used to produce a Rapid Evidence Summary (RES) for discussion with stakeholders. The summary comprised two research questions regarding innovations to address the screening backlog and what examples of best practice were available for effective and efficient use of cancer investigations, which could be continued beyond the pandemic.

### 1.1 Purpose of this review

The RES highlighted the impact of the pandemic on waiting times for endoscopic investigations for patients with potential Gastrointestinal (GI) cancers and that considerable work is needed to address the backlog and improve and recover detection rates and capacity (Rutter et al., 2020).

The RES provided evidence to show that, in order to prevent late-stage diagnosis, colorectal screening needs to be resumed as soon as possible (CADTH, 2020). Following the RES and discussion with the stakeholders, the decision was made to progress to rapid review to further explore the evidence regarding innovations which may address the backlog in endoscopic services and improve clinical outcome.

### 1.2 Research Question

| Review question |  |
| --- | --- |
| Which innovations can improve timeliness of investigations and address the backlog in endoscopy for patients with potential symptoms of GI cancers? |  |
| **Participants** | Patients over 18 years with potential symptoms GI cancer |
| **Intervention / exposure** | Cancer, Carcinoma, Adenocarcinoma, Neoplasms, Malignancy, Tumours Gastric, GI Oesophageal, Stomach, Bowel, Pancreatic, Pancreatobiliary, Colon, Rectal, Colorectal |
| **Comparison** | Colonoscopy, Endoscopy, Gastroscopy, Flexible sigmoidoscopy, Water assisted sigmoidoscopy, Oesophagogastroduodenoscopy; OGD, Endoscopic retrograde cholangiopancreatography; ERCP, Faecal immunochemical test (FIT) testing, CT colonography, Hepatico- |
|  | pancreatico-biliary endoscopic ultrasound, Capsule endoscopy, Device assisted enteroscopy, Barium swallow |
| **Outcomes** | Time to diagnosis, Stage at diagnosis, Reduction in waiting time, Diagnostic capacity, Triage, Reduction in backlog |
| **Other Study Considerations** |  |
| The pandemic was also considered with terms such as COVID-19 and pandemic |  |

## 2. RESULTS

### 2.1 Summary of the Evidence Base

Nine papers were identified for inclusion: seven studies and two reviews. Five studies reported the use of FIT to prioritise further testing, two studies reported interventions delivered via other settings such as primary care, these include cytopsponge testing and the Straight to Test (STT) pathway. The review evidence comprised a review on the novel colon capsule endoscopy (CCE) procedure and a summary of the effects of COVID-19 on CRC screening, the potential long outcomes, and ways to adapt CRC screening during the COVID-19 pandemic.

A narrative summary of the seven studies is provided below. The studies are presented by sub-headings based on the type of test used (e.g. cytosponge, FIT testing); and pathway adaptations/ innovations. The two reviews are presented narratively under the sub-headings of novel colon capsule endoscopy and individualized CRC screening for risk levels.

#### 2.1.1 Evidence from published studies

##### Cytosponge testing

di Pietro et al., (2020) conducted a study to examine whether Cytosponge testing could be used to triage patients referred for endoscopy for urgent investigations for oesophageal alarm symptoms.

In the study by di Pietro et al, (2020) of 123 patients referred to endoscopy at a single UK hospital, 21 patients were eligible for, and accepted Cytosponge testing. Incomplete swallow was reported in four patients, two of those patients were found to have glandular atypia and p53 positive cells suggestive of dysplasia or cancer. Another four patients had evidence of potential neoplasms. All eight of these patients were referred to endoscopy and four had cancer. A further three patients had cells suggestive of intestinal metaplasia and a diagnosis was made on two of the three. The remaining 10 patients had a normal Cytosponge result and were managed via telephone.

Thus, **Cytosponge may be useful in triaging patients with oesophageal symptoms**; however, the sample size in this feasibility study was small, and did not examine effectiveness of outcomes, which may be considered a significant limitation. Evaluation of clinical outcomes and whether adoption in primary care could increase the demand for upper GI endoscopy in the longer term is required.

##### FIT testing

In a letter to the British Journal of Surgery, Habib Bedwani et al., (2021) describe a single centre prospective study in which outstanding endoscopy patients from two week wait (2WW) referrals were given Faecal Immunochemical Testing (FIT) sampling kits for use at home.

The median wait for endoscopy was 87 days (95% CI 82 to 93 days). Of the 102 patients given tests, 66 patients completed FIT. Six patients were diagnosed with CRC. Only one patient diagnosed with CRC had completed FIT, which was raised. As this paper is a letter to the editor, no other statistical data are given.

This paper reports that the data from this study were used to create a model that may be used to triage patients referred for endoscopy during pandemic conditions. However, it also **reports that the model has not yet been implemented** and so there are no data on whether the model is acceptable, effective or efficient for use in practice. The letter is included as **policy makers may find the results of the model, when published, useful for decision-making surrounding CRC**.

Ho et al. (2021) used secondary data from nationally collected datasets regarding endoscopy services to conclude that **FIT testing could help to clear endoscopy backlogs caused by the COVID-19 pandemic**. FIT triaging of cases that are found to have greater than 10 µg haemoglobin per gram would reduce colonoscopy referrals to around 75% of usual levels. The authors suggested that by using FIT tests in this way, excluding those with <10 µg haemoglobin per gram from colonoscopy tests, could clear the backlog in endoscopy services in England by early 2022.

Loveday et al. (2020) evaluated the impact of **prioritising FIT to mitigate the impact of delays in the CRC urgent 2WW pathway**, which resulted from the COVID-19 pandemic. The study used modelling techniques for the 11,266 CRC patients in England diagnosed each year via the 2WW pathway. The study involved modelling FIT thresholds of 2,10 and 150µg Hb/g in the prioritisation of 2WW CRC referrals for colonoscopy. The outcomes estimated the reduction in survival and life years lost resulting from delays of 2- to 6-months for CRC patients in 2WW pathway.

The results of the modelling exercise found that delays in 2WW referral pathway are associated with substantial decreases in the 10-year survival of CRC patients. Delays of 2, 4 and 6 months across all 11,266 CRC patients per typical year via the 2WW pathway were estimated to result in 653, 1,4192 and 250 attributable deaths respectively and a loss of 9,214, 20,315 and 32,799 life years respectively. For stage 1 CRC patients aged 70 - 79, a 6-months delay was associated with a 12.5% reduction in survival. For stage 3 CRC patients for all age groups, a 2-months delay to surgery was associated with >9% reduction in survival and a 6-months delay was associated with >29% reduction in survival. For stage 3 CRC patients aged 30–39, a 4-month delay was associated with 9.4 life years lost and a six-month delay was associated with 15.1 life years lost. We note also, however, that there are uncertainties about the delays and influence on long-term disease-free survival (Garcia-Botello S et al, 2021).

Adopting a FIT threshold of 10µg Hb/g faeces could prioritise 18% of symptomatic CRC patients, avoid 89% of deaths attributable to presentational/diagnostic delay, and reduce immediate requirement for colonoscopy by >80%. Adopting a higher FIT threshold of 150µg Hb/g would prioritise 11% of symptomatic CRC patients, but at the expense of an additional 150, 326 and 517 deaths per year respectively if the delay rate was 2, 4 or 6 months when compared with the FIT threshold of 10µg Hb/g faeces.

In conclusion, there is evidence of better survival from prompt and prioritised referral to colonoscopy. **Potential symptomatic suspected cancer patients with a high FIT score should be prioritised for colonoscopy**. To avoid a substantial number of deaths, urgent attention is required to minimise and mitigate disruption to CRC diagnostics and treatment.

##### Adaption of a multilevel CRC intervention

Kruse-Diehr et al. (2021) conducted a case study of the development and adaption of a multilevel colorectal cancer (CRC) intervention at four primary care practices in rural Appalachian Kentucky, USA. One of the proposed study objectives was to conduct pilot testing of individually tailored evidence-based interventions that focus on CRC screening and follow-up; however, pilot testing was not performed as some of the study activities were temporarily halted due to the COVID-19 pandemic.

Researchers conducted a series of meetings with ‘clinic champions’ to discuss possible strategies to impact CRC screening rates based on new priorities following the COVID-19 pandemic. The authors do not describe whom the ‘clinic champions’ were in the paper.

Proposed strategies discussed by clinic champions included the **increased use of mailed stool-based testing, shifting from paper to digital educational tools and the increased use of telehealth to increase the number of annual wellness consultations** to promote CRC screening among patients to help increase CRC screening rates. Clinic champions highlighted the importance of moving away from a ‘colonoscopy first’ model of care to a ‘shared decision-making model’ whereby patients can decide to have an at-home stool-based test if they were dissatisfied with colonoscopy waiting times or the required COVID-19 testing before having the colonoscopy procedure. Moreover, clinic champions **advocated the use of the FIT-DNA test (i.e. Cologuard) over the FIT stool-based test as it uses fewer primary care clinic resources**. We note limited availability of Cologuard in UK currently, but which could be explored.

##### Pathway adaptations / innovations

Miller et al., (2021) report on a rapidly implemented ‘COVID-adapted triage pathway’ in Scotland, UK, whereby patients who were reporting with high-risk symptoms of CRC were triaged to quantitative faecal immunochemical tests (qFIT) + Computed Tomography (CT) colonography and the remainder underwent an initial qFIT to inform subsequent investigation.

Results reported similar cancer detection rates from the use of the ‘COVID-19 adapted triage pathway’ compared to a period before the pandemic; however, a 43% reduction in all primary care referrals was reported, so there may be selection bias in the comparison groups. The authors concluded that their **COVID-adapted triage pathway was successful in mitigating the adverse impacts on diagnostic capacity and could be a useful strategy to prevent delay to treatment while services remain limited** due to the COVID-19 pandemic.

Sagar et al. (2020) conducted an observational study of CRC patients managed via a Straight to Test (STT) pathway between 1 September 2019 and 19 March 2020 (pre-COVID-19 pandemic). In the STT pathway, patients proceed directly from GP review to specialist investigation without direct assessment by the secondary care team before investigation. In this study, the STT pathway was compared with a pre-STT pathway. 1,255 patients were referred onto the CRC pathway, with 43.7% (548) managed via STT and 56.3% (707) managed pre-STT by proceeding directly to face-to-face clinic consultation.

Of the 548 patients in the STT pathway, 56% were women, 44% were men. The average age was 68 years (range 20–79 years). The primary outcome measure was the proportion of patients who attained the 28-day diagnosis standard (i.e., the time at which the patient is informed whether they do or do not have CRC). The secondary outcome was the effect of the STT pathway on reducing face-to-face outpatient clinic appointments.

Prior to implementation of STT pathway, 82% of cancer referrals attained the 28-day target within a 7-month period (Jan 2019 to Jul 2019). For the pre-STT pathway, the colonoscopy rate was 68.6% and the CRC diagnosis rate was 6.4%. Following implementation of the STT pathway, 88% of cancer referrals attained the 28-day target in a 7-month period (Sep 2019 to Mar 2020). The colonoscopy rate was 81.5% and the CRC diagnosis rate was 7.1%. For patients on the STT pathway, the median time from referral to the patient being informed whether they did or did not have CRC was 14 days.

Assessment of the GP-led patient triage STT pathway found that 504 of 548 (92%) CRC clinic appointments were avoided. The GP assessment of patients agreed with the secondary care colorectal department in 93% of cases.

The findings of this study showed that the **GP-led patient triage STT pathway reduced time to diagnosis, evidenced by an improvement in attainment of the NHS 28-day Faster Diagnosis Standard**. In addition, the **STT pathway facilitated a reduction in face-to-face outpatient clinic appointments**, which could lead to a significant increase in available clinic time for more complex referrals.

Appropriateness as well as time taken through pathways is critical (Del Vecchio Blanco et al, 2021). Although this was a short study period of 7 months due to the onset of the pandemic in March 2020, **the GP-led patient triage STT pathway was accurate regarding patient suitability for colonoscopy and could have an important role to play in the early diagnosis of CRC**.

#### 2.1.2 Evidence from published reviews

##### Colon capsule endoscopy

A review paper by MacLeod et al., (2020) examines the novel **colon capsule endoscopy (CCE)** procedure, which is an innovative method for detecting colorectal pathology. While this is not a systematic review, it presents studies relating to the CCE procedure and discusses its relevance during the COVID-19 pandemic.

The review paper reports that CCE is a simple and safe procedure, with a low risk of adverse events that requires minimal training and can be conducted in the community by one healthcare professional, and therefore avoids the need for hospital attendance over the pandemic. **Published literature has shown CCE to be accurate for the detection of clinically significant polyps greater than 10mm in size** (Spada et al., 2016). Moreover, the review reports on a surveillance study whereby CCE was used as a triage test to colonoscopy and was found to reduce the number of colonoscopies by 43% (Kroijer at al., 2019). There are important implementation issues in terms of bowel preparation required and that require real-world evaluation (pilot work underway in Scotland).

Moreover, the review reports on a previous cost-effectiveness analysis, which assessed the additional cost of computerised tomography (CT) colonoscopy and CCE resulting from misdiagnoses in Canada (Palimaka et al., 2015). Palimaka et al., (2015) developed a deterministic Markov model to estimate the additional long-term costs and life-years lost due to false-negative results. The findings showed an additional cost of false-positive results for colon capsule endoscopy to be $0.41 per patient, while additional false-negatives for the CT colonoscopy arm generated an added cost of $116 per patient, with 0.0096 life-years lost per patient due to cancer. Palimaka et al., (2015) estimated this would result in an additional cost of $26,750 per life-year gained for colon capsule endoscopy compared with CT colonoscopy. The authors concluded that further research is needed to explore the cost implications of CCE compared to traditional colonoscopy with respect to the expected reductions in medical hardware costs and cost savings from the decrease in required colonoscopy procedures (MacLeod, Wilson and Watson, 2020). Furthermore, the review authors proposed that **CCE might be a potentially useful solution for investigating colorectal patients to reduce the demand on reinstated endoscopy units following the COVID-19 pandemic through effective triage**.

### Individualized CRC screening for risk levels

Kadakuntla et al. (2021) conducted a review summarising the effects of COVID-19 on CRC screening, the potential long outcomes, and ways to adapt CRC screening during the COVID-19 pandemic. Kadakuntla et al., (2021) state while the COVID-19 pandemic has been a major disruption to CRC screening, this may result in some beneficial changes to the current screening strategies. There may be a resulting shift from current CRC screening and surveillance practices towards the development of an individualized approach based on risk factors. There is potential to tailor FIT cut-off levels and for male/female etc. It will be different for true population screening, surveillance and symptomatic testing groups who are at different levels of pre-test risk (Cross et al, 2019). BSG Guidelines recommend those at highest risk remain on lists for investigations so FIT will be less useful for those groups (Rutter et al, 2020). Applying it to population screening could allow allocation of resources to people with higher risks and prevent inappropriate use of healthcare resources for those with lower risks.

### 2.2 Summary of the evidence base – included studies

**Table 1.** Summary of the identified studies.

| Study Reference | Methods and Setting | Key Outcomes | Comments |
| --- | --- | --- | --- |
| **Cytosponge testing** |  |  |  |
| di Pietro et al., (2020). | **Study Design**<br>Cytosponge is a non-endoscopic diagnostic tool consisting of a tethered capsule that is swallowed and collects immunohistochemical biomarkers of intestinal metaplasia and dysplasia. Cytosponge can be administered in primary or secondary care office settings.<br>This study assessed whether Cytosponge could be used to triage patients referred for urgent investigations of alarm oesophageal symptoms.<br>**Participants**<br>123 patients were referred for endoscopy, of which 21 were eligible for, and accepted Cytosponge. Mean age was 59.9 years.<br>**Outcome Measures**<br>The primary outcome measured was whether patients could be triaged to Cytosponge. | **Primary Findings**<br>Incomplete swallow was reported in 4 patients, 2 of those patients were found to have glandular atypia and p53 positive cells suggestive of dysplasia or cancer. Another 4 patients had evidence of potential neoplasms. All 8 of these patients were referred to endoscopy and 4 had cancer.<br>**Additional Findings**<br>A further 3 patients had cells suggestive of intestinal metaplasia and a diagnosis was made on 2 of the 3. The remaining 10 patients had a normal Cytosponge result and were managed via telephone. | This (feasibility) study shows that Cytosponge may be useful in triaging patients with oesophageal symptoms.<br>The study sample size is small and may be considered a significant limitation. |
| **FIT testing** |  |  |  |
| Habib Bedwani et al., (2021). | **Study Design**<br>Single-center prospective study in which all outstanding endoscopy from 2WW CRC referrals were given FIT sampling kits for home use. | **Primary Findings**<br>The median wait for endoscopy was 87 days (95% CI 82 to 93 days). 6 patients were diagnosed with CRC. Of the 102 patients given tests 66 patients completed FIT. Only 1 patient diagnosed with CRC had completed FIT. | This study reports how data from a single-center prospective study of FIT for 2WW referrals was used to create a model for triaging patients referred to endoscopy and potentially reducing the backlog. Long term results will follow the implementation of the model. |
|  | **Participants**<br>102 patients. Median age 61 years. 48% male; 52% female.<br>**Outcome Measures**<br>Outcomes included time to diagnostics, cancer yield, uptake of FIT and results of tests. Data from this study was used to create a prioritisation model for endoscopy and CRC 2WW referrals during the C-19 pandemic. | The FIT result was significantly higher in the presence of neoplasia. Using this data, the study team have created the prioritisation model for 2WW referrals. | Further research may be needed to test the model. |
| Ho et al., (2021). | **Study Design**<br>A secondary national analysis of data for colonoscopies, flexible sigmoidoscopies, and gastroscopies from National Health Service (NHS) Trusts in NHS England's Monthly Diagnostic Waiting Times and Activity dataset.<br>**Participants**<br>Patients who received colonoscopies, flexible sigmoidoscopies, gastroscopies, and total endoscopic procedures in 125 NHS England trusts.<br>**Outcomes Measures**<br>The primary outcomes were the change in number of endoscopic procedures compared with the same month in 2019, and an estimate of the backlog of procedures associated with the pandemic.<br>Secondary outcomes were the effect of increasing capacity, a temporary reduction of capacity, and the effect of FIT triaging on the backlog of procedures. | **Primary Findings**<br>FIT triaging of cases for patients with greater than 10 µg haemoglobin per gram would reduce colonoscopy referrals to around 75% of usual levels, with the backlog cleared in early 2022. | A national analysis of collected data sets in NHS England suggest that FIT triaging would help to clear endoscopy backlogs caused by the COVID-19 pandemic. |
| Loveday et al., (2021). | **Study Design**<br>To evaluate the impact of prioritising faecal immunochemical testing (FIT) to mitigate the impact of delays in the colorectal cancer (CRC) urgent (2-week-wait (2WW)) pathway which resulted from the COVID-19 pandemic. | **Primary Findings**<br>Delays of 2/4/6 months across all 11,266 CRC patients per typical year via the 2WW pathway were estimated to result in 653/1,419/2,250 attributable deaths and a loss of 9,214/20,315/32,799 life years.<br>Adopting an FIT threshold of 10µg Hb/g faeces prioritized out of | Delays in 2WW referral pathway delays are associated with substantial decreases in the 10-year survival of CRC patients.<br>Short delays in CRC 2WW pathway from diagnosis to treatment could result in significant mortality and lost life years, and thus a gain in survival from prompt investigatory referral to colonoscopy. |
|  | **Participants**<br>Modelling for the 11,266 CRC patients diagnosed per year via the 2WW pathway. Modelling of FIT thresholds of 2, 10 and 150µg Hb/g in the prioritization of 2WW referrals for colonoscopy.<br>**Outcome Measures**<br>Reduction in survival and life years lost resulting from delays of 2- to 6-months for CRC patients in 2WW pathway.<br>Comparing FIT thresholds of 2, 10 and 150µg Hb/g to prioritise 2WW referrals for colonoscopy. | delay 18% of symptomatic CRC patients, avoided 89% of deaths attributable to presentational/diagnostic delay while reducing immediate colonoscopy requirement by >80%.<br>Adopting a higher FIT threshold of 150µg Hb/g prioritized out of delay 11% of symptomatic CRC patients, but at the expense of an additional 150/326/517 deaths per year if the delay rate was 2/4/6 months when compared with FIT threshold of 10µg Hb/g faeces.<br>**Additional Findings**<br>For stage 1 CRC patients aged 70 - 79, a 6-month delay was associated with a 12.5% reduction in survival.<br>For stage 3 CRC patients for all age groups, a 2-month delay to surgery was associated with >9% reduction in survival and a 6-month delay was associated with >29% reduction in survival.<br>For stage 3 CRC patients aged 30–39, a 4-month delay was associated with 9.4 life years lost and a six-month delay was associated with 15.1 life years lost. | FIT-positive CRC patients should be prioritized out of delay for colonoscopy.<br>To avoid substantial numbers of deaths, urgent attention is required to minimise and mitigate disruption to CRC diagnostics and treatment.<br>One limitation is that with any model-based analysis, the accuracy of estimation is dependent on the validity of assumptions and estimates used for parameterisation.<br>Evaluation is required about whether most colonoscopy will still be required and the overall reduction in demand may be less substantial. (Ed: pilot data in Aneurin Bevan UHB are consistent with this) |
| **Adaption of a multilevel CRC intervention** |  |  |  |
| Kruse-Diehr, et al., (2021). | **Study Design**<br>Case study. Development and adaptation of a Multilevel Colorectal Cancer (CRC) screening Project in four rural primary care practices in Appalachian Kentucky, USA. The study had a one-year project duration.<br>**Participants**<br>Meetings were held with clinic champions to discuss implementation of strategies to impact clinic CRC screening rates.<br>**Outcome Measures** | **Primary Findings**<br>Clinic champions were more open to increasing mailed, homebased stool-based testing efforts as a screening option.<br>Clinic champions were interested in exploring novel interventions to increase screening recommendations and annual wellness visits to assist in recovery of CRC screening rates.<br>**Additional Findings**<br>Strategies such as shifting from paper to digital educational tools and using telehealth | Meetings with clinic champions highlighted new priorities following the pandemic.<br>The study findings offer screening strategies following discussions with clinic champions.<br>The study does not provide explicit information/description of clinic champions.<br>Discussions were prioritised to focus on how clinics can change from a “colonoscopy first” model to a |
|  | Proposal of strategies to impact CRC screening dates following the pandemic. | to increase annual wellness visits for screening were reported.<br>Clinics were eager to consider any evidence-based interventions (EBIs) that could help elevate screening to pre-pandemic levels.<br>Greater interest in education on communication methods to motivate patients to get screened was discussed. | shared decision-making model in which patients could choose at-home stool-based options if they were not comfortable with colonoscopy wait times or COVID testing requirements prior to colonoscopy.<br>The pilot testing of individually- tailored, multilevel evidence-based interventions that address CRC screening and follow-up were not conducted due to the study being paused due to the pandemic. |
| **Pathway adaptations / innovations** |  |  |  |
| Miller et al., (2021). | **Study Design**<br>Diagnostic test accuracy study in Scotland for patients entering pathway between 1/4/20 and 31/5/20.<br>**Participants**<br>422 (median age 64 years, 220 women). Patients referred with high-risk symptoms of lower gastrointestinal disease. Patients reporting high-risk symptoms triaged to quantitative faecal immunochemical tests (qFIT) +CT. The remainder triaged to qFIT to inform subsequent investigation.<br>**Outcome Measures**<br>Cancer detection rates during the COVID-adapted triage pathway compared with cancer detection rates pre-pandemic. | **Primary Findings**<br>202 patients triaged to CT and qFIT (47.9%); 211 (50.0%) to qFIT only, eight (1.9%) to outpatient clinic and one to colonoscopy.<br>13 cancers were detected (3.1%), similar to the mean cancer detection rate from all referrals in 2017–2019 (3.3%).<br>Compared with the period 1 April–31 May in 2017–2019, a 43% reduction in all primary care referrals was observed (1071 referrals expected reducing to 609). | COVID-adapted pathway mitigated the adverse effects on diagnostic capacity and detected cancer at the expected rate within those referred.<br>The overall reduction in the number of referrals was substantial. The described risk mitigating measures could be a useful addition whilst standard diagnostic services remain constrained due to the ongoing pandemic.<br>Prioritization of symptomatic patients through the use of FIT and CT minimal preparation scans has been shown to be a rational approach to mitigate risk and prevent delay to treatment when access to endoscopy is limited. (Ed: NB however, different FIT thresholds exist between Scotland (80 mcg/g), England (120 mcg/g) and Wales (150 mcg/g) – so the numbers and proportions of screen detected vs symptomatic cancers is consequently likely to be different.) |
| Sagar et al., (2021). | **Study Design**<br>Observational study of CRC patients managed via a Straight to Test (STT) pathway from 1/9/20 to 19/3/20.<br>**Participants**<br>In STT pathway, patients proceed directly from GP review to specialist investigation without | **Primary Findings**<br>STT CRC pathway improves attainment of NHS England 28-day Faster Diagnosis Standard and the effect of the pathway on reducing face-to-face outpatient clinic appointments.<br><u>Prior to implementation of STT pathway:</u> | An STT CRC pathway with GP-led patient triage reduces time to diagnosis, evidenced by an improvement in attainment of the NHS 28-day Faster Diagnosis Standard, and facilitates reduction of face-to-face outpatient clinic appointments which can lead to significant increase in available clinic time for expedited review of more complex referrals.<br>One limitation of this study is the short study period of 7 months. |
|  | direct assessment by the secondary care team prior to investigation.<br>In this study, STT pathway was compared with pre-STT pathway.<br>1255 patients were referred on the CRC pathway, with 43.7% (548) managed via STT and 56.3% (707) proceeded directly to face-to-face clinic consultation.<br>Of the 548 patients in STT pathway, 56% were women, 44% were men. The average age was 68 years (range 20–79 years).<br>**Outcome Measures**<br>The primary outcome was the proportion of patients who attained the 28-day diagnosis standard (time at which the patient is informed whether they do or do not have CRC). The secondary outcome was the effect of the STT pathway on reducing face-to-face outpatient clinic appointments. | 82% of cancer referrals attained 28-day target within a 7-month period (Jan 2019 to Jul 2019)<br>Colonoscopy rate of 68.6% and CRC diagnosis rate of 6.4%<br><u>Following implementation of STT pathway:</u><br>88% of cancer referrals attained 28-day target in 7-month period (Sep 2019 to Mar 2020).<br>Colonoscopy rate of 81.5% and a CRC diagnosis rate of 7.1%.<br>For patients on the STT pathway, the median time from referral to the patient being informed whether they did or did not have CRC was 14 days.<br><u>Assessment of GP-led triage</u><br>Of 548 STT referrals, 504 (92%) CRC clinic appointments were avoided.<br>GP assessment of patients agreed with colorectal department in 93% of cases. | GP-led triage regarding patient suitability for colonoscopy is accurate and has an important role in the CRC pathway. |

#### Summary of the evidence base – included reviews

**Table 2.** Summary of the identified reviews.

| Citation | Included studies | Inclusion criteria | Quality | Results | Observations/Notes |
| --- | --- | --- | --- | --- | --- |
| **Colon capsule endoscopy** |  |  |  |  |  |
| MacLeod et al., (2020). | The review paper does not provide details on the number of included studies, study design or search dates. | Studies relating to the relevance of the colon capsule endoscopy during the COVID-19 pandemic were discussed in the review paper. Explicit inclusion criteria were not provided in the review paper. | This was not a systematic review paper. Quality appraisal of the included papers was not performed. | Faecal immunochemistry tests (FITs) are valuable methods to prioritise patients waiting for further investigation during the coronavirus pandemic.<br>FITs can be carried out in the community and have a high level of accuracy.<br>CT of the abdomen and pelvis has been proposed as a triage tool for use over the pandemic and allows patients to be prioritised for colonoscopy once services resume. CT carries risk of radiation and increases footfall of patients at hospital settings.<br>Colon capsule endoscopy (CCE) is an innovative technology for visualizing the colon which can be carried out in the community, reducing hospital attendance, and can be carried out by one healthcare professional.<br>Published literature has shown CCE to be accurate for the detection of clinically significant polyps greater than 10mm in size (Spada et al., 2016).<br>Recent surveillance study has used CCE as a filter to colonoscopy and demonstrated a reduction in the number of colonoscopies required (Kroijer et al., 2019). | This review paper is not a primary source of research but offers insights into the novel CCE procedure with respect to COVID-19 considerations. This review paper provides an overview of the literature relating to the relevance of CCE during the COVID-19 pandemic and its ability to triage patients effectively to further endoscopic investigations. The review concludes that CCE is a promising technology which could be used to reduce colonoscopy demand and effectively triage patients to colonoscopy with appropriate urgency. |

| Individualized CRC screening for risk levels |  |  |  |  |  |
| --- | --- | --- | --- | --- | --- |
| Kadakuntla et al., (2021). | This is a literature review paper and does not provide details on the number of included studies, study design or search dates. | This review includes studies of the effects of COVID-19 on Colorectal Cancer (CRC) screening, the potential long outcomes, and ways to adapt Colorectal Cancer screening during the pandemic. Specific inclusion criteria are not provided. | Quality appraisal of included studies was not performed. | FIT may reduce the need for colonoscopies by up to 80%. FIT is also relatively inexpensive and less invasive than colonoscopy. The use of telehealth may increase accessibility and facilitate implementation of FIT. Follow ups can also be managed via text messages and phone calls.<br>The backlog caused by COVID-19 may also be addressed via open access colonoscopies, this may also reduce costs by avoiding pre procedure consultations. | This is a literature review and as such provides no primary data. However, it does present some valuable data from other studies on how the backlog in colonoscopy may be addressed and costs potentially reduced. |

### 2.3 Bottom line results

The review found five studies relating to the use of FIT testing. The evidence suggests that increased use of FIT testing could reduce the endoscopy backlog and save NHS resources. However, it was not possible to draw robust conclusions about the relative value for money of increased use of FIT testing in Wales.

Two studies explored triage in primary care settings such as GP surgeries suggest that using innovations such as the cytosponge for oesophageal symptoms and/or direct referral to specialist investigation could reduce backlog and speed up time to diagnosis.

The two included reviews suggest that the FIT test may be a good way forward in terms of CRC screening during the COVID-19 pandemic and could potentially triage patients effectively to further endoscopic investigations. One of the included reviews concluded that CCE is a promising technology that could be used to reduce colonoscopy demand and effectively triage patients to colonoscopy with appropriate urgency.

## 3. DISCUSSION

### 3.1 Summary

The review found the most evidence relating to FIT testing, with 5 studies identified in the review. (Habib Bedwani et al., 2021; Ho et al., 2021; Kruse-Diehr, et al., 2021; Loveday et al., 2021; Miller et al., 2021).

Kadakuntla et al., (2021) and MacLeod et al., (2020) suggest from the findings of their reviews that the FIT test may be a good way forward in terms of CRC screening during the COVID-19 pandemic and could potentially triage patients effectively to further endoscopic investigations.

There was evidence that primary care settings could be used to triage patients using innovations such as the cytosponge for oesophageal symptoms (di Pietro et al., 2020). Sagar et al., (2021) suggest patients could proceed directly from GP review to specialist investigation without direct assessment by the secondary care team before investigation. This could reduce backlog and speed up time to diagnosis (Sagar et al., 2021).

### 3.2 Implications for policy and practice

This review highlights four main areas that policy makers may wish to explore further, these are:

1. Increased implementation of FIT testing in the symptomatic referral patient cohort in Primary care.
2. Increased use of the results of the FIT test for prioritising patients for further diagnostic testing in secondary care
3. Exploring potential effects of direct referral from the GP to specialist investigations.

Considering increased FIT testing, the Welsh National Framework for the Implementation of FIT in the Symptomatic Service (NHS Wales, 2021) states the following:

*“In 2017 NICE published ‘Quantitative faecal immunochemical tests to guide referral for colorectal cancer in primary care’ (DG30). This guidance recommended the use of FIT to guide referral for people without rectal bleeding who have unexplained symptoms but do not meet the criteria for a suspected cancer pathway referral outlined in NICE’s guideline NG12 ‘Suspected cancer: recognition and referral’. In February 2019 Health Technology Wales (HTW) reviewed the evidence and published a report (HTW Guidance 007) which supported the adoption of DG30 in Welsh Health Boards, stating that “NHS Wales should adopt this guidance or justify why it has not been followed”. In order to ensure standardised implementation of DG30 and to evaluate emerging data to inform future policy on the use of FIT in other cohorts such as the NG12 suspected cancer group, a [National Endoscopy Programme] FIT subgroup was established in 2020 to develop the agreed national framework”*

Thus, the policy implications of increased FIT testing reflect current national framework guidance.

### 3.3 Limitations of the available evidence and quality statements

The review only found eight studies and two reviews that were relevant to the review questions. The review did not identify any randomised controlled trials (RCTs) or evidence to support the cost-effectiveness of innovations. However, these study designs often taken years to achieve results, and this was a rapid review to find current available evidence.

The included evidence is recently published, with small samples (di Pietro et al., 2020), short-term follow up periods (Sagar et al., 2020) and assumptions required for modelling (Loveday et al., 2021). This may reduce the generalisability and confidence of conclusions.

#### Quality statements

The confidence in the strength of evidence about FIT testing was rated as ‘low-moderate confidence’.

Some included studies performed well against the quality criteria set by the corresponding JBI Checklist (n=5). However, there were cases where the included study methods employed were not as rigorous (n=2) or well described as the checklist required (n=3).

The key limitations of included studies and reviews are summarised in this review. However, due to the rapid nature of this review no formal risk of bias was conducted.

### 3.4 Strengths and limitations of this Rapid Review

This rapid review was designed to answer a specific question as part of the Wales COVID-19 Evidence Centre Work Programme. This question was developed in consultation with a stakeholder, Professor Tom Crosby OBE. Professor Crosby is a Consultant Oncologist, National Cancer Clinical Director for Wales and Clinical Lead for Transforming Cancer Services.

This was a rapid review, but every effort was made to conduct the review with as much rigour as possible within the timeframe. The main strengths and limitations are described below.

#### Strengths

The review team took a rigorous and systematic approach. The team liaised with the stakeholder, Professor Tom Crosby, regarding the research question and search strategy. The review team used expert feedback from information scientists and members of the Wales COVID-19 Evidence Centre to check the search strategy. The search strategy was tested in two electronic databases, before being used in all databases.

Two reviewers undertook screening of titles and abstracts and completed full text screening. Both screening stages were conducted using predefined inclusion/ exclusion criteria. In cases of disagreement, these were discussed, and an agreement was reached by both reviewers.

A single reviewer conducted the extraction and quality appraisal, with (25%) of the included literature checked by a second reviewer.

The review has identified recently published literature with recommendations that could help to address the backlog in endoscopy for patients with potential symptoms of GI cancers.

#### Limitations

The timing and speed of the review meant that the review team could only find published literature.

The team also wishes to highlight that one of the included pieces of literature by Habib Bedwani et al (2021), discusses a model they are evaluating, but evaluation of this model is yet to be published, and the paper did not assess its potential usefulness in answering the research question.

The research question itself is built upon innovations, which may be difficult to identify depending upon keywords chosen by authors and how the evidence is indexed in electronic repositories. The review team mitigated this risk as much as possible by seeking expert opinion on the search strategy and testing the search strategy before use in the review.

Full integration/ synthesis of review findings could not be achieved due to heterogeneity of studies; therefore, a narrative synthesis was used.

Key limitations of included studies and reviews are summarised. However, due to the rapid nature of this review no formal risk of bias was conducted.

## Data Availability

All data produced in the present study are available upon reasonable request to the authors

## RAPID REVIEW METHODS

The literature search was conducted in June 2021. Initial search terms were developed by the review team and then supplemented by adding terms and key words from previously published studies that were relevant to this review. The terms were checked by an information scientist and two colleagues from the Wales COVID-19 Evidence Centre who have substantial experience in systematic reviewing.

### 5.1 Eligibility criteria

The scope and eligibility criteria for the rapid review are shown in Table 3. This review includes all GI cancers and all investigations for their detection. The scope is limited to OECD countries with healthcare systems comparable to the UK. The scope is limited to UK settings due to the NHS context, but no other geographic limits have been applied. All study designs are eligible for inclusion in the review.

**Table 3.** Scope and eligibility criteria.

|  | Inclusion | Exclusion |
| --- | --- | --- |
| **Participants** | All adults having endoscopic investigations or investigations for symptoms of GI cancers screened for gastrointestinal cancer | Participants younger than 18 years or not having endoscopic investigations or investigations for potential GI cancer symptoms. Asymptomatic adults not screened for gastrointestinal cancer |
| **Intervention/exposure** | Screening Investigations for potential for gastrointestinal cancer |  |
| **Comparator** | New interventions screening devices/services<br>Shifting of investigations screening from one setting to another (e.g. secondary care to primary care) |  |
| **Outcome measures** | Stage of cancer at diagnosis<br>Waiting times for referral/diagnosis |  |
| **Setting** | UK NHS OECD countries |  |
| **Study design** | Any study design will be considered (RCT, cohort, observational) |  |
| **Study quality** | Study quality will be graded |  |
| **Other factors**<br>*Any other key points to note* | The study area is gastrointestinal cancer, and therefore we will be looking at all types of endoscopies and all gastrointestinal cancers<br>English or Welsh language studies only<br>Published since 2016 |  |

### 5.2 Literature search strategy

The literature search was conducted in June 2021. Key sources listed below were searched for full text published papers, published between 2016-2021, to find the most up to date published evidence. The searches were limited to published research in the English or Welsh languages, and within the UK context. Unpublished or non-full text work and grey literature will not be included in the review. The scope outlined for this search is to keep the review concise and deliverable within the timeframe expected of a rapid review.

### 5.3 Resources List

The search strategy for Medline is presented below [also noting the platform used, e.g., OVID] (which has been approved by the stakeholders) or key words for COVID-resources. The strategy was adapted for use across other platforms.

| Key Sources | Search strategy |
| --- | --- |
| 1. Medline | 1. Endoscopy/ |
| 2. Embase | 2. Endoscopy.ti,ab |
| 3. ASSIA | 3. 1 OR 2 |
| 4. CINAHL | 4. Waiting lists/ |
| 5. Cochrane Library | 5. Capacity building/ |
| 6. HTA libraries | 6. Waiting time* or waiting list* or delay* or capacity or efficiency or productivity |
| 7. MEDLINE | 7. 4 OR 5 OR 6 |
| 8. Prospero | 8. COVID-19/ |
|  | 9. COVID OR pandemic.ti,ab |
|  | 10. 8 OR 9 |
|  | 11. 3 AND 7 AND 10 |
|  | 12. Limit to 2020 to present time |

### 5.4 Study selection process

Results from the literature searches were imported into Covidence reference management system, where duplicates were removed. Title and abstracts were screened for inclusion followed by full text screening. Both screening stages were undertaken by two reviewers against predefined inclusion/exclusion criteria. In cases of disagreement, these were discussed, and an agreement was reached by both reviewers.

### 5.5 Study selection flow chart

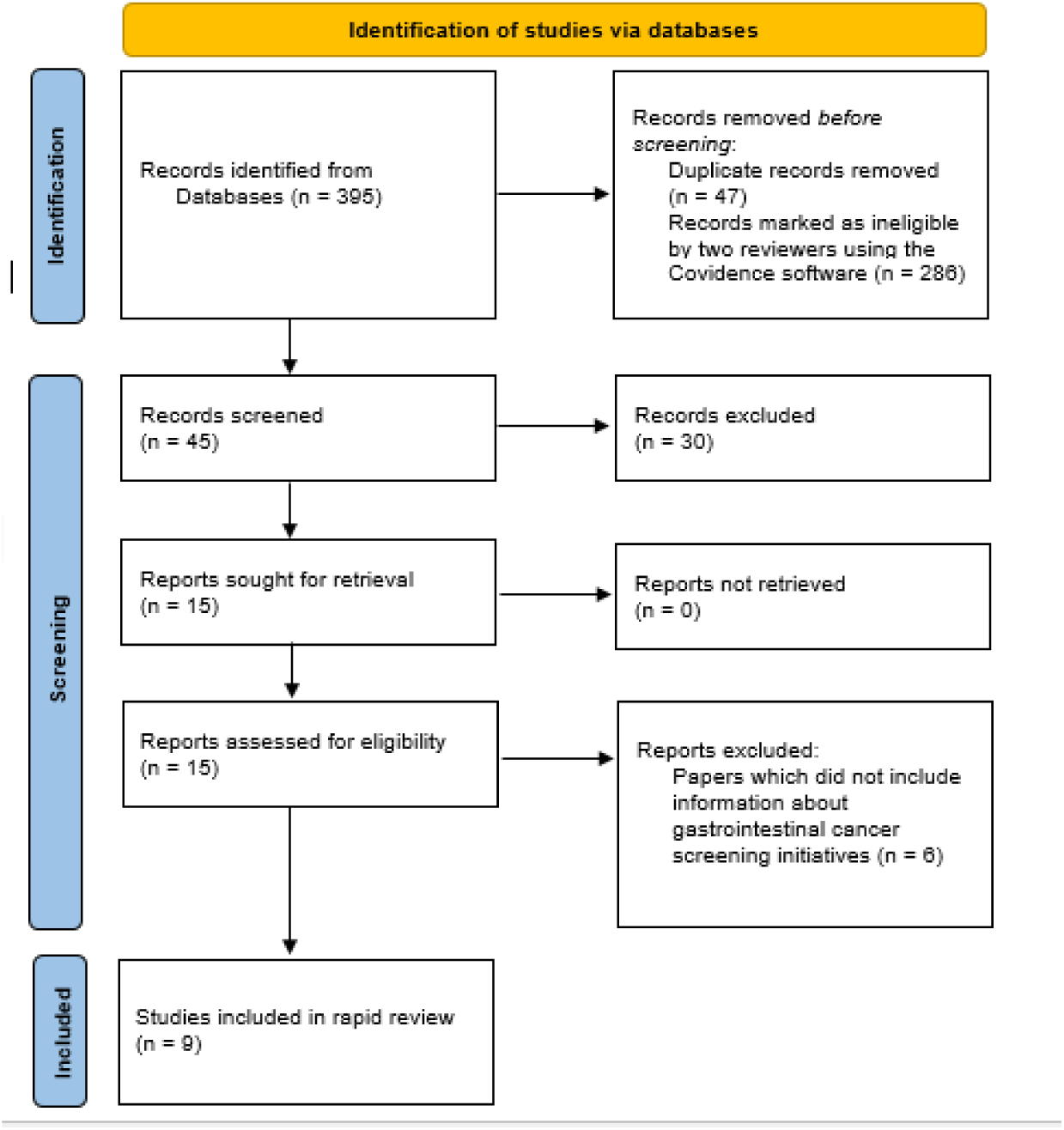

*From:* Page MJ, McKenzie JE, Bossuyt PM, Boutron I, Hoffmann TC, Mulrow CD, et al. The PRISMA 2020 statement: an updated guideline for reporting systematic reviews. doi: https://doi.org/10.1136/bmj.n71

### 5.6 Data extraction

A standardised data extraction table was created and performed by a single reviewer, with a quarter of extractions checked by a second reviewer.

The following information was extracted for all studies when reported:

- study citation (author, year of publication)
- study details (study design, geographical region, data collection dates)
- study participants (sample size, type of participant: i.e. doctor, nurse, mixed HSCW etc.)
- study outcomes
- study results (Baseline characteristics etc.)
- additional notes

### 5.7 Quality appraisal

Quality appraisal was carried out by a single reviewer, using; the JBI analytical cross-sectional study checklist; the JBI systematic reviews and research syntheses checklist; the JBI case reports checklist; and the JBI cohort studies checklist.

A quarter of quality appraisals were checked by a second reviewer Discrepancies arising during quality appraisal were discussed and an agreement was reached by reviewers.

The checklists were assigned to the included studies and reviews by the review team. The decision was based on choosing the most appropriate quality appraisal tool from the methods described in the included papers.

Tables 4-7 report the findings of the quality appraisals. Y indicates Yes; N indicates No; N/A indicates Not Applicable; and UC indicates Unclear.

**Table 4.** JBI analytical cross-sectional study checklist.

| Citation | Q1. Were the criteria for inclusion in the sample clearly defined? | Q2. Were the study subjects and the setting described in detail? | Q3. Was the exposure measured in a valid and reliable way? | Q4. Were objective, standard criteria used for measurement of the condition? | Q5. Were confounding factors identified? | Q6. Were strategies to deal with confounding factors stated? | Q7. Were the outcomes measured in a valid and reliable way? | Q8. Was appropriate statistical analysis used? |
| --- | --- | --- | --- | --- | --- | --- | --- | --- |
| di Pietro et al., (2020). | N | N | UC | UC | N | N | UC | UC |
| Habib Bedwani et al., (2021). | N | N | N | N | N | N | N | N |
| Ho et al., (2021). | Y | Y | Y | Y | Y | Y | Y | Y |
| Loveday et al., (2021). | Y | Y | Y | Y | N | N | Y | Y |
| Sagar et al., (2021). | Y | Y | Y | Y | N | N | Y | Y |

**Table 5.**
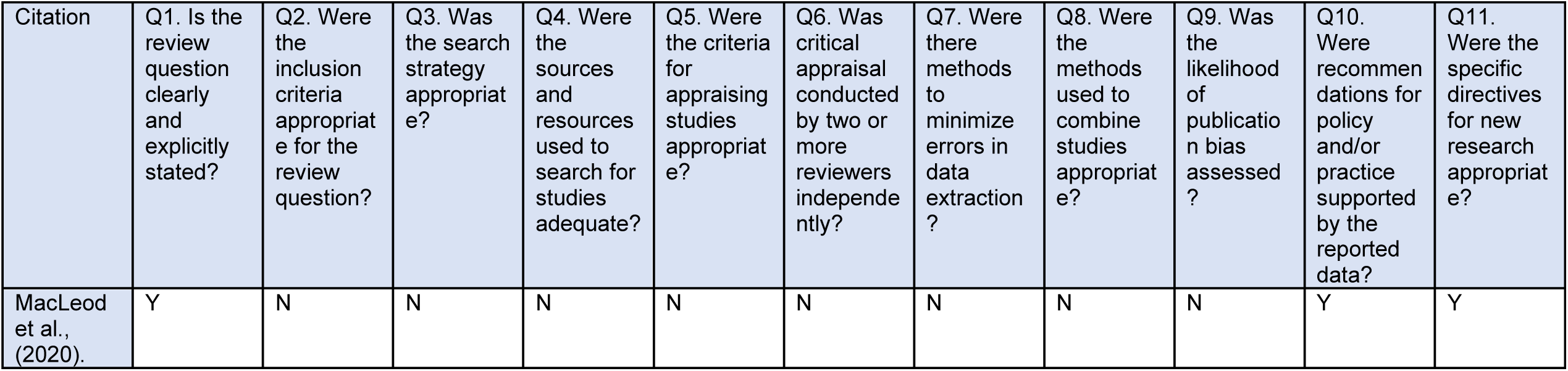
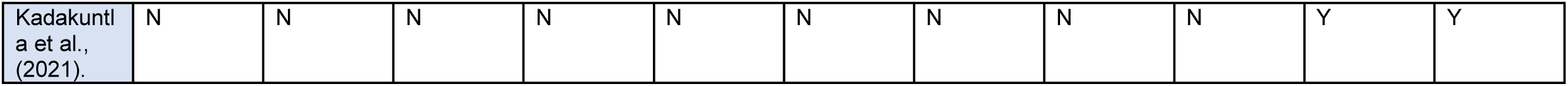
JBI Systematic Reviews and Research Syntheses Checklist.

| Citation | Q1. Is the review question clearly and explicitly stated? | Q2. Were the inclusion criteria appropriate for the review question? | Q3. Was the search strategy appropriate? | Q4. Were the sources and resources used to search for studies adequate? | Q5. Were the criteria for appraising studies appropriate? | Q6. Was critical appraisal conducted by two or more reviewers independently? | Q7. Were there methods to minimize errors in data extraction? | Q8. Were the methods used to combine studies appropriate? | Q9. Was the likelihood of publication bias assessed? | Q10. Were recommendations for policy and/or practice supported by the reported data? | Q11. Were the specific directives for new research appropriate? |
| --- | --- | --- | --- | --- | --- | --- | --- | --- | --- | --- | --- |
| MacLeod et al., (2020). | Y | N | N | N | N | N | N | N | N | Y | Y |
| Kadakuntla et al., (2021). | N | N | N | N | N | N | N | N | N | Y | Y |

**Table 6.** JBI Critical Appraisal Checklist for Case Reports.

| Citation | Q1. Were patient's demographic characteristics clearly described? | Q2. Was the patient's history clearly described and presented as a timeline? | Q3. Was the current clinical condition of the patient on presentation clearly described? | Q4. Were diagnostic tests or assessment methods and the results clearly described? | Q5. Was the intervention(s) or treatment procedure(s) clearly described? | Q6. Was the post-intervention clinical condition clearly described? | Q7. Were adverse events (harms) or unanticipated events identified and described? | Q8. Does the case report provide takeaway lessons? |
| --- | --- | --- | --- | --- | --- | --- | --- | --- |
| Kruse-Diehr et al., (2021). | N | N | N | N | N | N | N | Y |

**Table 7.**
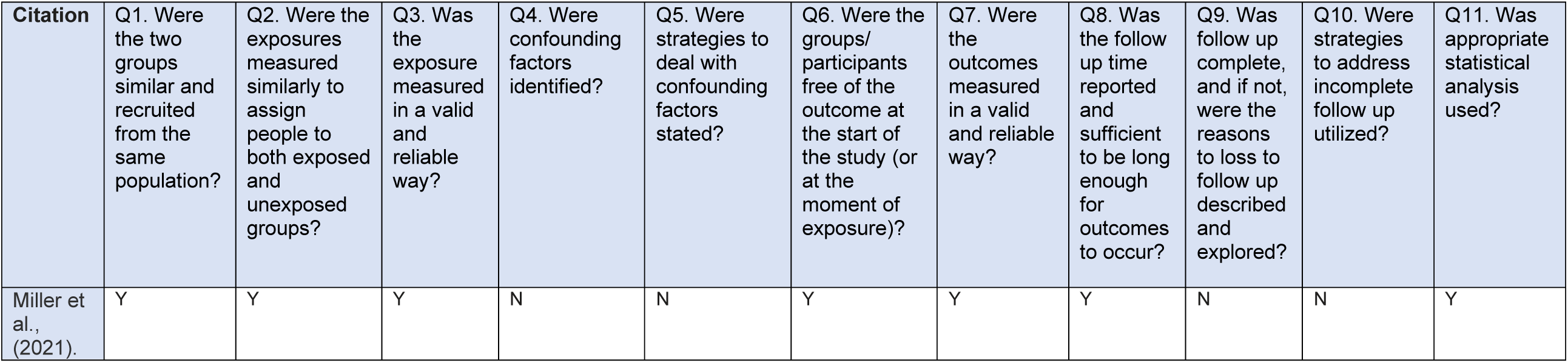
JBI Critical Appraisal Checklist for Cohort Studies.

| Citation | Q1. Were the two groups similar and recruited from the same population? | Q2. Were the exposures measured similarly to assign people to both exposed and unexposed groups? | Q3. Was the exposure measured in a valid and reliable way? | Q4. Were confounding factors identified? | Q5. Were strategies to deal with confounding factors stated? | Q6. Were the groups/ participants free of the outcome at the start of the study (or at the moment of exposure)? | Q7. Were the outcomes measured in a valid and reliable way? | Q8. Was the follow up time reported and sufficient to be long enough for outcomes to occur? | Q9. Was follow up complete, and if not, were the reasons to loss to follow up described and explored? | Q10. Were strategies to address incomplete follow up utilized? | Q11. Was appropriate statistical analysis used? |
| --- | --- | --- | --- | --- | --- | --- | --- | --- | --- | --- | --- |
| Miller et al., (2021). | Y | Y | Y | N | N | Y | Y | Y | N | N | Y |

### 5.8 Synthesis

The findings of this review are presented narratively. Data from the included studies are summarized and presented in Tables 1 and 2 in section 2.

## 6.2 Acknowledgements and author contributions

The authors would like to thank Professor Tom Crosby for acting as expert stakeholder; Dr Ruth Lewis and Dr Alison Cooper for providing support to the review team; Dr Elizabeth Gillen for reviewing our search strategy and conducting the searches in EMBASE; Ms Maggie Hendry for reviewing our search strategy and providing valuable feedback; Professor Deborah Fitzsimmons for reviewing the report and providing critical feedback. Dr Sunil Dolwani for external peer review.

## Author contributions

Project leads: CW, RTE, NB; drafting of report: JMC, RTE; contribution to writing and critical editing of the report; JMC, BA, NH, AH, JR, LHS, NB, CW, RTE; reviewing: JMC, BA, NH, AH, JR, LHS

## 6.3 Abbreviations

Acronym: Full Description
2WW: pathway 2-week-wait pathway
ADR: Adenoma detection rate
AEC: Ambulatory endoscopy centre
CCE: procedure Colon capsule endoscopy procedure
CRC: Colorectal Cancer
CRUK: Cancer Research UK
CT: colonography Computed Tomography colonography
DES: Discrete Event Simulation
FIT: Faecal immunochemical testing
FS: Flexible Sigmoidoscopy
GI: cancers Gastrointestinal cancers
GP: General Practitioner
JBI: Joanna Briggs Institute
NCRI: National Cancer Research Institute
NHS: National Health Service
PHE: Public Health England
PICO: framework Participant, Intervention, Comparison, Outcomes framework
qFIT: Quantitative faecal immunochemical tests
RCT: Randomised Controlled Trial
RES: Rapid Evidence Summary
STT: pathway Straight to Test pathway
WCEC: Wales COVID-19 Evidence Centre

## 7 ABOUT THE WALES COVID-19 EVIDENCE CENTRE (WCEC)

The WCEC integrates with worldwide efforts to synthesise and mobilise knowledge from research.

We operate with a core team as part of Health and Care Research Wales, are hosted in the [Wales Centre for Primary and Emergency Care Research (PRIME)](http://www.primecentre.wales/), and are led by [Professor Adrian Edwards of Cardiff University](http://www.primecentre.wales/).

The core team of the centre works closely with collaborating partners in [Health Technology Wales](https://www.healthtechnology.wales/), Wales Centre for Evidence-Based Care, [Specialist Unit for Review Evidence centre](https://www.cardiff.ac.uk/specialist-unit-for-review-evidence), [SAIL Databank](https://healthandcareresearchwales.org/about-research-community/secure-anonymised-information-linkage-sail-databank), [Bangor Institute for Health & Medical Research](https://www.bangor.ac.uk/health-sciences/research/index.php.en), Health and Care Economics Cymru and the [Public Health Wales Observatory](https://phw.nhs.wales/services-and-teams/observatory/).

Together we aim to provide around 50 reviews per year, answering the priority questions for policy and practice in Wales as we meet the demands of the pandemic and its impacts.

### Director

Professor Adrian Edwards

### Website

https://healthandcareresearchwales.org/about-research-community/wales-covid-19-evidence-centre

